# Overt and occult hypoxemia in patients hospitalized with novel coronavirus disease 2019

**DOI:** 10.1101/2022.06.14.22276166

**Authors:** Shrirang M. Gadrey, Piyus Mohanty, Sean P. Haughey, Beck A. Jacobsen, Kira J. Dubester, Katherine M. Webb, Rebecca L. Kowalski, Jessica J. Dreicer, Robert T. Andris, Matthew T. Clark, Christopher C. Moore, Andre Holder, Rishi Kamaleswaran, Sarah J. Ratcliffe, J. Randall Moorman

## Abstract

**Background:** Progressive hypoxemia is the predominant mode of deterioration in COVID-19. Among hypoxemia measures, the ratio of the partial pressure of arterial oxygen to the fraction of inspired oxygen (P/F ratio) has optimal construct validity but poor availability because it requires arterial blood sampling. Pulse oximetry reports oxygenation continuously, but occult hypoxemia can occur in Black patients because the technique is affected by skin color. Oxygen dissociation curves allow non-invasive estimation of P/F ratios (ePFR) but this approach remains unproven.

**Research Question:** Can ePFRs measure overt and occult hypoxemia?

**Study Design and methods:** We retrospectively studied COVID-19 hospital encounters (n=5319) at two academic centers (University of Virginia [UVA] and Emory University). We measured primary outcomes (death or ICU transfer within 24 hours), ePFR, conventional hypoxemia measures, baseline predictors (age, sex, race, comorbidity), and acute predictors (National Early Warning Score (NEWS) and Sepsis-3). We updated predictors every 15 minutes. We assessed predictive validity using adjusted odds ratios (AOR) and area under receiver operating characteristics curves (AUROC). We quantified disparities (Black vs non-Black) in empirical cumulative distributions using the Kolmogorov-Smirnov (K-S) two-sample test.

**Results:** Overt hypoxemia (low ePFR) predicted bad outcomes (AOR for a 100-point ePFR drop: 2.7 [UVA]; 1.7 [Emory]; p<0.01) with better discrimination (AUROC: 0.76 [UVA]; 0.71 [Emory]) than NEWS (AUROC: 0.70 [UVA]; 0.70 [Emory]) or Sepsis-3 (AUROC: 0.68 [UVA]; 0.65 [Emory]). We found racial differences consistent with occult hypoxemia. Black patients had better apparent oxygenation (K-S distance: 0.17 [both sites]; p<0.01) but, for comparable ePFRs, worse outcomes than other patients (AOR: 2.2 [UVA]; 1.2 [Emory], p<0.01).

**Interpretation:** The ePFR was a valid measure of overt hypoxemia. In COVID-19, it may outperform multi-organ dysfunction models like NEWS and Sepsis-3. By accounting for biased oximetry as well as clinicians’ real-time responses to it (supplemental oxygen adjustment), ePFRs may enable statistical modelling of racial disparities in outcomes attributable to occult hypoxemia.

---

Modelling the risk of adverse outcomes from novel Coronavirus disease 2019 (COVID-19) has been an area of intense investigation. Two recent systematic reviews identified over 200 new models, nearly half of which modeled risk of adverse outcomes (clinical deterioration, critical illness, or mortality)^1,2^. We reviewed the predictors that were reported as being useful in these reviews and seven subsequent studies^3–9^. Since progressive hypoxemia is the predominant mode of deterioration in COVID-19, we expected hypoxemia markers to be the strongest predictors. Hypoxemia markers, however, predicted outcomes in only 7 models (<10%)^3,6,8,10–13^. This points to an opportunity to improve the hypoxemia markers used in clinical practice and research.

The most commonly featured hypoxemia markers were the oxygen saturation of binding sites of hemoglobin from pulse oximetry (SpO_2_, %) and the oxygen flow rate (liters per minute). Most models only used SpO_2_, without regard to oxygen supplementation^3,10–12,14^. This approach loses power when patients with differing oxygen supplementation levels are compared (Figure 1; Scenarios 2, 3, 5). It is also affected by practice patterns like SpO_2_ targets and promptness of weaning supplemental oxygen. The National Early Warning Score (NEWS) models include oxygen supplementation, but in a binary form where 2 points are assigned for supplemental oxygen use, regardless of the flow rate. The resulting scores do not always reflect severity of hypoxemia (Figure 1; Scenarios 2, 3, and 5).

**Figure 1:**
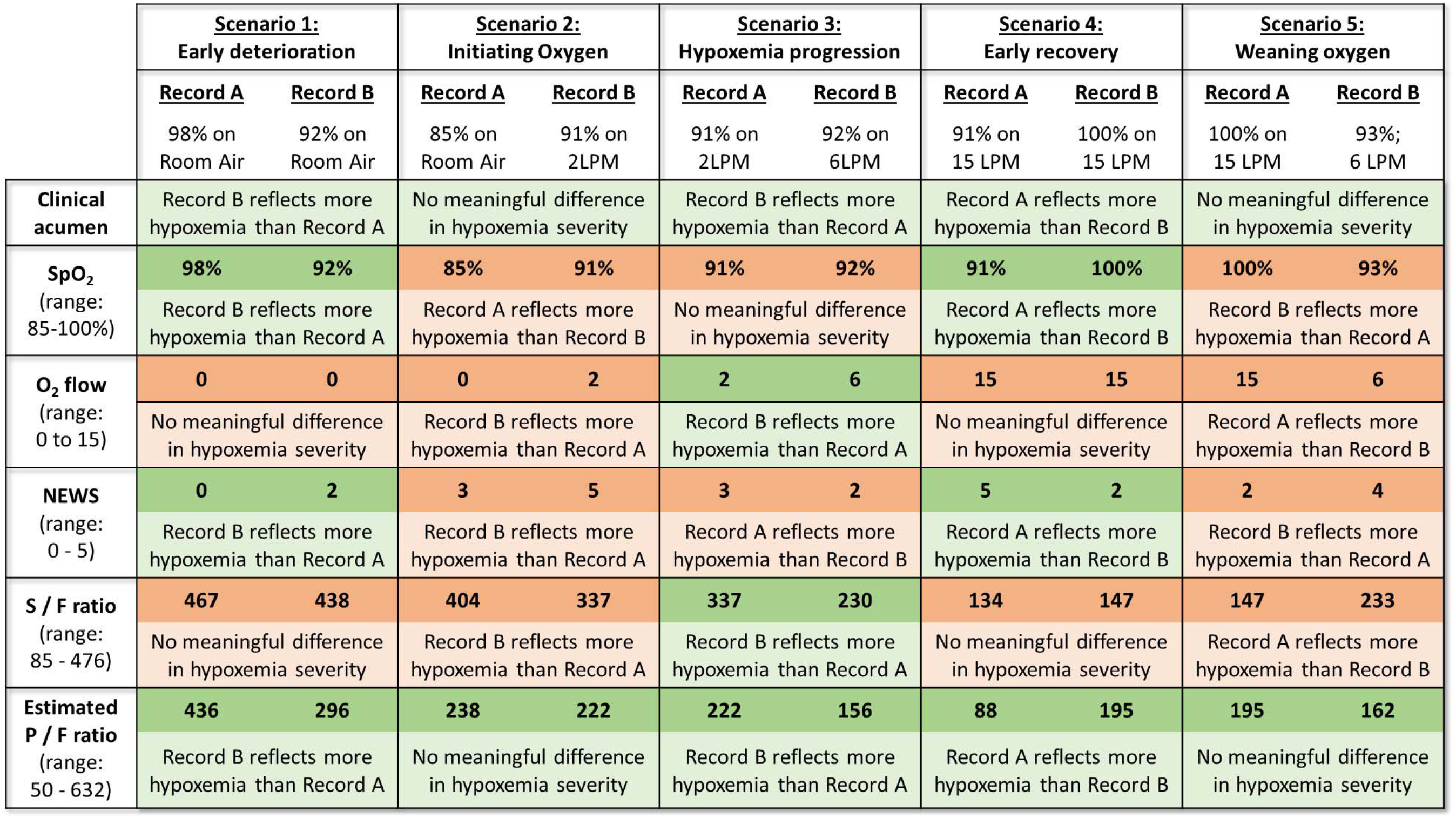
Evaluation of the construct validity of operational markers of hypoxemia in hypothetical clinical scenarios. Construct validity of any marker of hypoxemia is the extent to which that marker accurately reflects the clinical construct of hypoxemia. This figure examines the construct validity of five operational markers of hypoxemia (rows) in common clinical scenarios (columns). In each scenario (column), two records of a patient’s oxygenation are compared (Record A on left, Record B on right). The first row titled “clinical acumen” describes a clinically sensible conclusion that a clinician might draw by comparing the two records. For example, in Scenario 2, a clinician will likely conclude that the two records do not represent any meaningful change in the severity of hypoxemic respiratory failure (row 1, column 2). Rather, Record B (SpO_2_ of 91% on 2LPM of oxygen) might simply reflect the fact that a clinician initiated supplemental oxygen in response to Record A (SpO_2_ of 85% on room air). Each of the subsequent rows describes the conclusion based solely on comparing a particular marker of hypoxemia. For example, if one solely compared SpO_2_ in Scenario 2 (row 2, column 2), the conclusion would be that Record A reflects significantly more severe hypoxemia than Record B (SpO_2_ of 85% v/s 91%). Considering the varying range of each marker, we used the following cutoffs to determine a “significantly more/less hypoxemia”: any difference ≥ 1 for NEWS (range 0 to 5), any difference ≥ 2 for SpO_2_ (range 85 to 100) and supplemental oxygen flow rate (range 0 to 15 LPM), and any difference ≥ 50 for S/F ratio (range 85 - 476) and P/F ratio (range 50 - 632). A cell is shaded green when there is agreement between the marker of hypoxemia and clinical acumen; and it is shaded red when there is disagreement. This figure illustrates the advantages of estimated P/F ratios over other markers – it is the only marker to agree with clinical acumen in all scenarios. We were unable to conceptualize any scenario where P/F ratio would be inferior to other markers. (RA = Room Air; LPM = liters per minute)

The ratio of the partial pressure of arterial oxygen (PaO_2_, mm Hg) to the fraction of inspired oxygen (FiO_2_, no units), the P/F ratio, does not suffer from these drawbacks. We found only two models that include it – Sepsis-3 and Toward a COVID-19 Score (TACS)^13,15^. In both cases, PaO_2_ is measured on arterial blood gas (ABG) samples. When PaO_2_ was unavailable, the Sepsis-3 researchers used multiple imputation with chained equations and the TACS researchers imputed a P/F ratio of 381 (assuming PaO_2_ at 80 and FiO_2_ at 0.21 [room air]). However, as the proportion of missing data increases, these imputation methods become increasingly more unreliable^16^. Outside the intensive care unit (ICU), ABGs are missing in over 75% of cases^17,18^. It is not surprising, therefore, that the only models that used the measured P/F ratio were derived in the ICU.

The ratio of the SpO_2_ to the FiO_2_, the S/F ratio, has been used^7^, but its construct validity is limited. The SpO_2_ range (typically 85-100%) is narrower than the corresponding PaO_2_ range (50 to 130 mmHg). Thus, FiO_2_ settings play a larger role in the S/F ratio than in the P/F ratio (Figure 1: rows 3 and 5 agree in all scenarios, rows 3 and 6 do not). Additionally, the S/F ratio sidesteps the fact that the relationship between PaO_2_ and SpO_2_ is not a straight line. Judging hypoxemia severity using S/F ratios can, therefore, be misleading (Figure 1; Scenarios 1, 2, 4, and 5).

To allow non-invasive estimation of P/F ratios, we derived the following oxygen dissociation curve model from a cohort of hospitalized, non-intubated patients with simultaneous ABG and pulse oximetry recordings^19^:

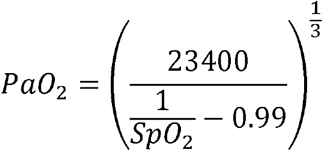

Older models were derived from laboratory solutions of hemoglobin^20,21^ or whole blood specimens of a few young, healthy males^22^. They underestimated the severity of hypoxemia when applied to hospitalized patients. The newer model remedied this drawback^19^. The P/F ratios estimated using this model (ePFRs) have high construct validity in all scenarios (Figure 1). We hypothesized that ePFRs are a valid measure of overt hypoxemia. If so, clinicians might use the ubiquitous SpO_2_ to monitor ePFRs continuously without being limited by arterial blood draws.

The relationship between pulse oximetry saturation (SpO_2_) readings and arterial oxygen saturation (SaO_2_) is complicated. Pulse oximetry often overestimates arterial oxygenation, especially in darker skinned individuals^23–31^. One study showed a racial bias in pulse oximetry readings which led to “occult hypoxemia” (undiagnosed arterial desaturation) at three times the frequency in Black patients as compared to White patients^25^. Another study showed that even in the absence of bias, occult hypoxemia was more frequent among darker skinned individuals due to a lower precision of oximetry readings^26^. Occult hypoxemia may have deleterious effects on outcomes of darker skinned individuals.

The ideal method to model the impact of occult hypoxemia on outcomes is unclear. Comparisons between simultaneously recorded SpO_2_ (pulse oximetry) and SaO_2_ (ABG) are limited by their exclusion of the majority of patients in whom arterial blood sampling is unavailable. Studying the population-wide distributions of SpO_2_ may not be an appropriate alternative because these distributions are influenced by clinicians’ real time efforts to maintain SpO_2_ in a particular range (typically 90-94%) by adjusting patients’ supplemental oxygen settings. The ePFR overcomes this barrier by simultaneously accounting for any falsely reassuring pulse oximetry readings (the corresponding PaO_2_ estimate) as well as clinicians’ real-time responses to that false reassurance (lower FiO_2_ setting). We therefore hypothesized that comparing population-wide distributions of ePFR by race would reveal occult hypoxemia and allow better modelling of its impact on clinical outcomes.

## Study Design and Methods

### 2.1) Data collection

We identified a retrospective cohort of adults (age ≥ 18 years) with hospital encounters (emergency room visit and/or hospital admission) for acute COVID-19 at the University of Virginia Medical Center (UVA), an academic tertiary-care center. We identified 1172 instances where the first positive SARS-CoV-2 test occurred in the context of a hospital encounter. Only the first positive test was used. We excluded: (a) 9 encounters that lacked any vitals, tests or notes; (b) 17 encounters where chart reviews showed that the timing of the SARS-CoV-2 infection did not match the hospital encounter (usually patients whose first positive test in our record was deemed to be a persistently positive test after a resolved infection at another facility); (c) 46 encounters where the ICU admission and/or mortality occurred within 4 hours of encounter start time (which was necessary in the primary analysis because we censored data 4 hours prior to time of outcome). The final cohort consisted of 1100 encounters in the first year of UVA’s pandemic experience (March 2020 to February 2021).

To ensure reproducibility of findings in diverse populations, we studied similar encounters at two hospitals affiliated with the Emory University: the Emory University Hospital (EUH) and Emory University Hospital Midtown (EUH-M). While UVA serves a rural and predominantly White population, the Emory sites serve an urban and predominantly Black population. While UVA and EUH are university hospitals, EUH-M is a community-based academic hospital. The Emory sites had 12,784 COVID-19 hospital encounters by December 2021. We randomly sampled a third of these encounters (n = 4219). This ensured that the Emory dataset represented more phases of the pandemic than the UVA dataset.

At UVA, we manually reviewed all charts to (a) confirm acute COVID-19, (b) separate pre-infection baseline SOFA from acute SOFA (eTable 1), and (c) ascertain the Charlson Comorbidity Index (CCI, eTable 2). Seven of the authors (JD, KD, SG, SH, BJ, RK, and KW) were the reviewers. This procedure was not repeated in the Emory data.

We queried the data warehouse to record (a) baseline risk predictors (age, sex, race, height, weight, Charlson comorbidity index), (b) all components of ePFR, S/F ratio, SOFA score, and NEWS (eTable 3; eFigure 1), and (c) the time of transfer to ICU and/or death. We updated predictors every 15 minutes from encounter start time. In the absence of new data, nursing flow sheet variables (e.g. vital signs, mental state assessments, and supplemental oxygen settings) were carried forward for 12 hours (the typical nursing shift) and laboratory values were carried forward for 24 hours (the typical frequency of phlebotomy in acute illness). We censored data four hours prior to the time of outcome.

To ensure adequate inter-rater reliability, we followed best practices including clear operational definitions and standardized abstraction forms^32^. For data entry, we used REDCap hosted at the University of Virginia^33,34^. To measure inter-rater reliability (IRR), we randomly sampled 10% of each reviewer’s charts and conducted blinded second reviews on those charts. Our IRR metrics were percent agreement and Krippendorf’s alpha^35^. We pre-specified adequate reliability as (a) alpha ≥ 0.8 or (b) 0.8 > alpha ≥ 0.67 with agreement ≥ 90%.

### 2.2) Characterizing the risk of clinical deterioration associated with overt hypoxemia

The primary outcome of interest was clinical deterioration, defined as transfer to an ICU or in-hospital mortality. We validated the ePFR as a measure of overt hypoxemia in two ways. First, we calculated adjusted odds ratio (AOR; logistic regression) to determine the extent to which ePFR was associated with clinical deterioration after adjusting for all non-hypoxemia components of the NEWS and Sepsis-3 (SOFA) models (temperature, heart rate, respiratory rate, mean arterial pressure, Glasgow Coma Scale, creatinine, platelet count, total bilirubin). In this model, we used all variables in a continuous form, rather than the categorical form prescribed in NEWS and SOFA.

Second, we measured the rise in area under receiver operating characteristic curves (AUROC) when the ePFR was added to a baseline risk model. The baseline risk model included age, sex, race, and Charlson Comorbidity Index. At UVA, the baseline risk model additionally included the baseline SOFA from chart reviews. For comparison, we measured the rise in AUROC associated with addition of conventional hypoxemia measures (SpO_2_, oxygen flow rate, and S/F ratio), and multi-system organ dysfunction scores (NEWS and SOFA) to the same baseline model. We used the Delong test to compare AUROCs^36^.

### 2.3) Characterizing racial disparities attributable to occult hypoxemia from pulse oximetry

To characterize the influence of skin color on predictive validity of pulse oximetry based hypoxemia measures (ePFR, S/F ratio, and SpO_2_), we used race as a surrogate for skin color, and compared patients whose medical records indicate their race to be Black with all other patients^23,25^. We computed empirical cumulative distribution functions (ECDFs) for each measure and quantified racial differences (Black vs Non-Black) in the distributions using the Kolmogorov-Smirnov (K-S) two sample test. We also visualized, by race, the relationship between the hypoxemia measure and the risk of imminent clinical deterioration. We used AOR (logistic regression) to quantify the influence of race on this relationship.

### 2.4) Sensitivity analyses in UVA data and other details

We tested the impact of restricted cubic splines (3 knots) for temperature, heart rate, respiratory rate, and mean arterial pressure, since either extreme of these vital signs are associated with clinical deterioration. We assessed the impact on the estimated predictive validity of the ePFR of excluding the patients who (a) died without transfer to ICU (b) who were discharged without outcome in < 24 hours (most likely to be emergency room visits and brief observation stays). For our primary analysis we used a prediction horizon of 24 hours. We repeated the analysis for 3, 5, 7, and 14 day horizons. We also varied the censoring from 4 hours before to the time of outcome in a secondary analysis. In all our regression models, we used the Huber-White method for robust standard error to correct for correlation from repeated measures.

We used R version 3.5.1 to perform all analyses^37^. The University of Virginia and Emory Institutional Review Boards approved the study (Protocol 20249 at UVA; STUDY-00000302 at Emory).

## Results

### 3.1) Cohort characteristics and inter-rater reliability of chart reviews

At UVA, we analyzed 399,797 every-15-minute rows (1100 individuals). The primary outcome occurred in 177 (17%) patients. At Emory, we analyzed 1,510,070 every-15-minute rows (4219 individuals). The primary outcome occurred in 791 (19%) patients. The probability that a random row was followed by the outcome within 24 hours was 1.9% at UVA and 2.9% at Emory. The demographic and clinical cohort characteristics are outlined in Table 1.

**Table 1:** UVA cohort Emory cohort.

| Clinical Variable | UVA cohort |  | Emory cohort |  |
| --- | --- | --- | --- | --- |
|  | All | Outcome | All | Outcome |
|  | patients | positive | patients | positive |
|  | (1100) | (177) | (4219) | (791) |
| Age in years, median (IQR) | 55 (38-68) | 67 (57-77) | 55 (39-68) | 64 (52-75) |
| Male, n (%) | 545 (50) | 101 (57) | 2016 (48) | 453 (57) |
| Race/Ethnicity, n (%) |  |  |  |  |
| White, Non-Hispanic | 446 (40) | 89 (50) | 987 (23) | 215 (27) |
| Black | 320 (29) | 54 (31) | 2515 (60) | 422 (54) |
| Hispanic | 285 (26) | 31 (17) | 327 (8) | 64 (8) |
| Other | 49 (5) | 3 (2) | 390 (9) | 90 (11) |
| Charlson Comorbidity Index, n (%) |  |  |  |  |
| 0 | 526 (48) | 49 (28) | 1713 (41) | 116 (15) |
| 1-2 | 299 (27) | 55 (31) | 1701 (40) | 320 (40) |
| ≥ 3 | 275 (25) | 73 (41) | 805 (19) | 355 (45) |
| Baseline SOFA, n (%) |  |  |  |  |
| 0 | 722 (66) | 76 (43) |  | NA |
| 1-2 | 271 (24) | 63 (36) |  | NA |
| ≥ 3 | 107 (10) | 38 (21) |  | NA |
| In-hospital mortality, n (%) | 49 (5) | 49 (28) | 240 (6) | 240 (30) |
| ICU Transfer, n (%) | 161 (15) | 161 (91) | 694 (16) | 694 (88) |
| Composite Outcome, n (%) | 177 (17) | 177 (100) | 791 (19) | 791 (100) |

Of the manually abstracted data, agreement was 79% for CCI and 95% for baseline SOFA; alpha was 0.84 for CCI and 0.90 for baseline SOFA. This met our pre-specified inter-rater reliability threshold.

### 3.2) Risk of clinical deterioration associated with overt hypoxemia

Overt hypoxemia, operationalized using ePFR, independently predicted clinical deterioration within 24 hours (AOR: 0.990 [UVA], 0.995 [Emory]). Adding ePFR to the baseline risk model resulted in model discrimination (AUROC: 0.76 [UVA]; 0.71 [Emory]) that was better than SpO_2_ (AUROC: 0.65 [UVA]; 0.66 [Emory]), oxygen flow rate (AUROC: 0.73 [UVA]; 0.69 [Emory]) and comparable to S/F ratio (AUROC: 0.76 [UVA]; 0.70 [Emory]). At both sites, ePFR outperformed NEWS (AUROC: 0.70 [UVA]; 0.70 [Emory]) and Sepsis-3 (AUROC: 0.68 [UVA]; 0.65 [Emory]) (Figure 2).

**Figure 2:**
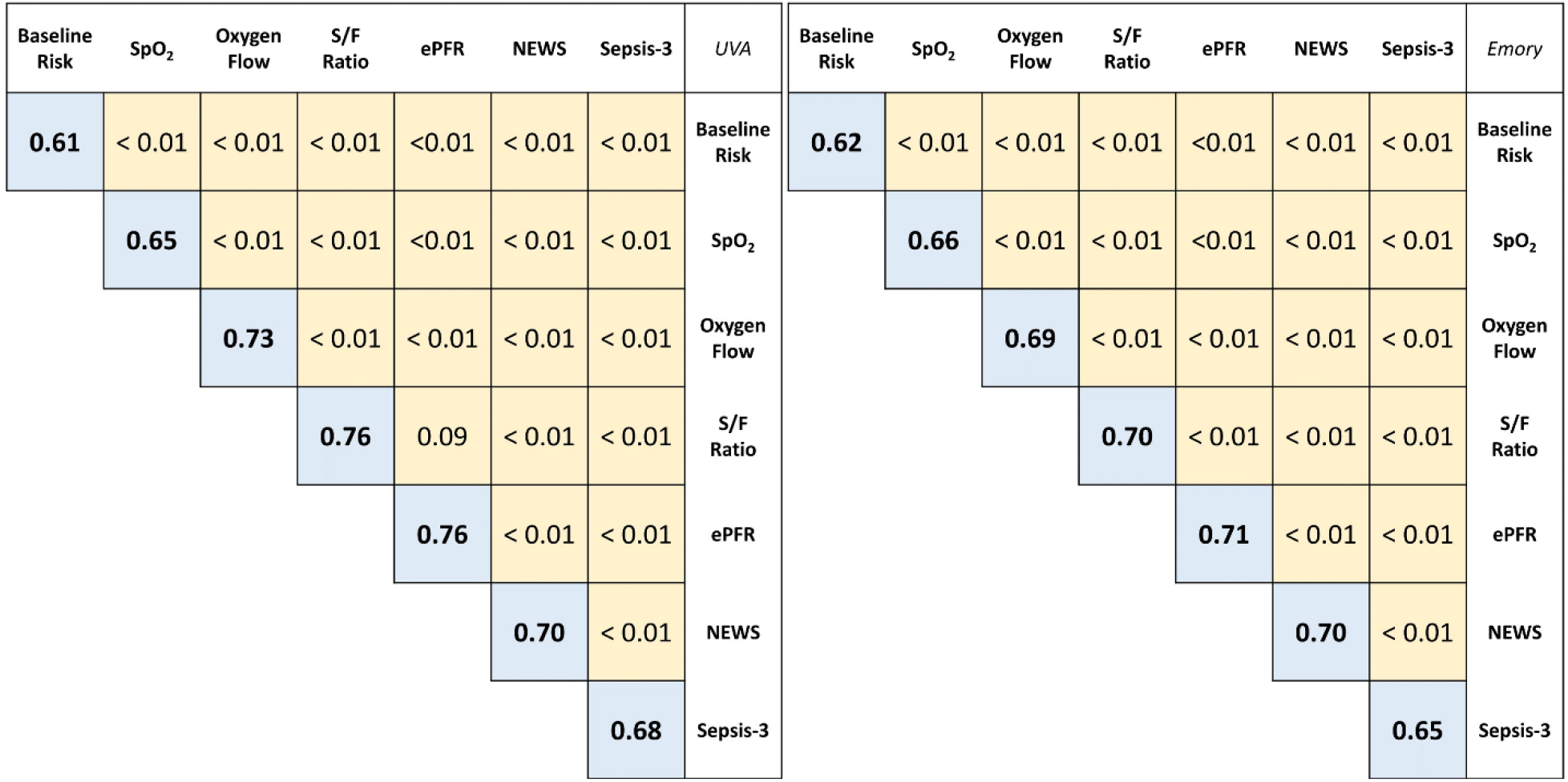
Discrimination of estimated P/F ratio for clinical deterioration in patients with COVID-19. This figure compares the Area Under the Receiver Operator Characteristic curve (AUROC) of multivariable logistic regression models for clinical deterioration (transfer to ICU or mortality within 24 hours) from COVID-19. The blue boxes show the AUROC for a model and the yellow boxes show p-values from pairwise comparison (DeLong’s test). Results from UVA are on the left and those from Emory are on the right. The baseline risk model used age, sex, race, Charlson Comorbidity Index, and pre-infection baseline Sequential Organ Failure Assessment (SOFA) score as predictors (baseline SOFA was only available at UVA). The model for the each criterion was created by adding that criterion to the baseline risk predictors. The estimated P/F ratio (ePFR) had optimal model discrimination, and it outperformed NEWS and Sepsis-3 (acute rise in SOFA score at UVA and total SOFA in Emory) models.

### 3.3) Racial disparities attributable to occult hypoxemia from pulse oximetry

For all 3 hypoxemia measures (SpO_2_, S/F ratio, and ePFR) and for both sites (UVA and Emory), we observed that the ECDF were “right-shifted” in Black patients relative to non-Black patients; that is, Black patients appeared to have better oxygenation (higher SpO_2_, S/F ratios and ePFR) than non-Black patients. Yet, Black patients had worse outcomes for comparable degrees of apparent oxygenation (Figure 3 and eFigure 2).

**Figure 3:**
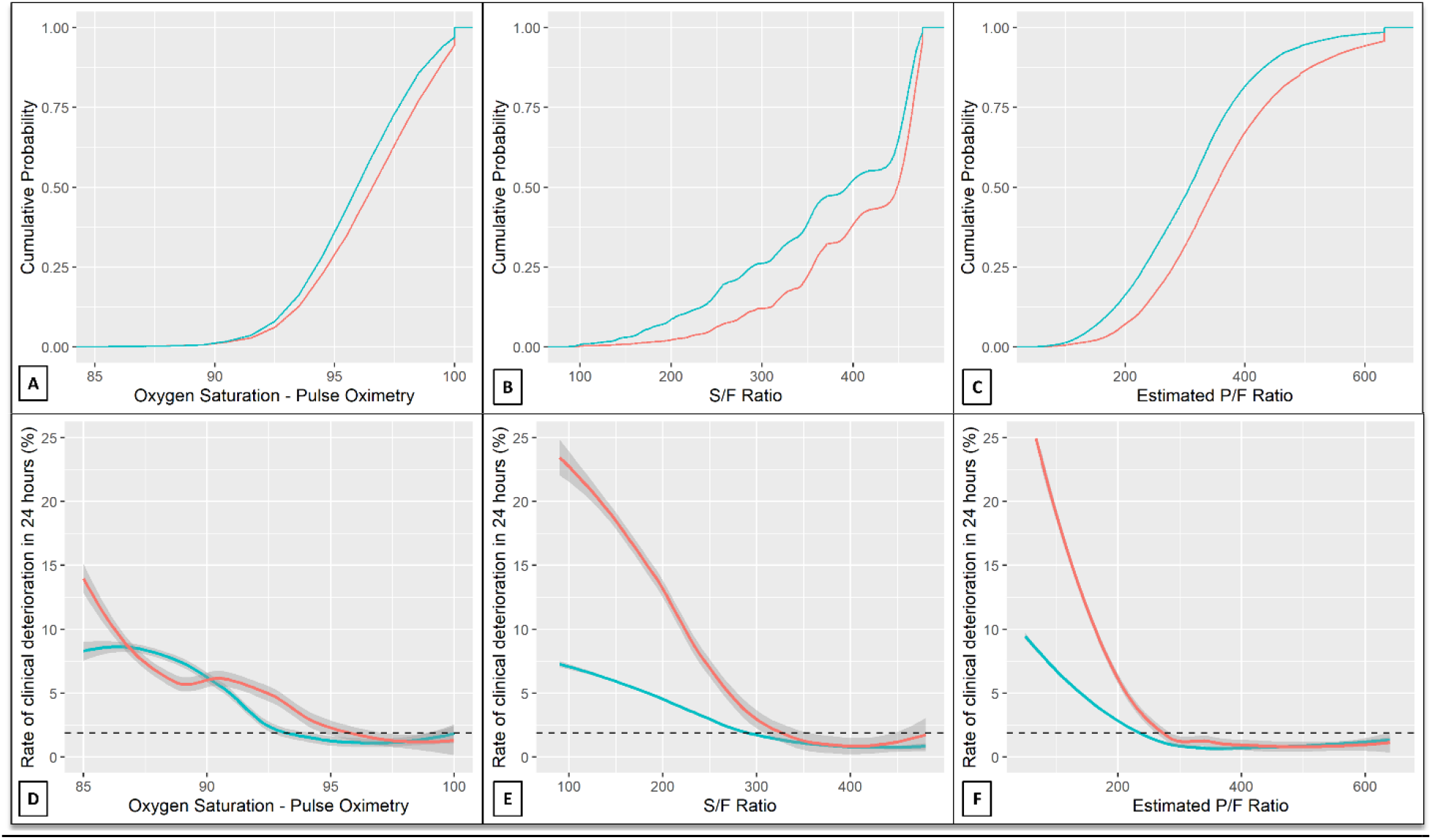
Characterizing the impact of racially biased pulse oximetry measurements. Panels A - C show the Empirical Cumulative Distribution Functions for SpO_2_, S/F ratio, and ePFR respectively. This figure depicts the results from UVA. Corresponding results from Emory are shown in eFigure 2. Race is encoded by color (red - Black patients, blue - others). The separation in SpO_2_ distributions was narrow (being minimal at SpO_2_ < 92%), suggesting an equitable clinician effort to prevent oxygen desaturation. Yet, the separation in S/F ratio and ePFR distributions was wide at all values. This suggests that, on average, clinicians were achieving their SpO_2_ targets with lower FiO_2_ settings in Black patients (eFigure 4). For comparable S/F ratio and ePFR values, outcomes were worse for Black patients than others (Panel E-F). Together, these findings reveal that clinicians were likely undertreating hypoxemia due to an overestimation of SpO_2_. Significantly, this disparity remained undetected when the SpO_2_ was studied instead of S/F ratio or ePFR (Panel D). To make the plots directly comparable despite the varying scales of the hypoxemia measures, we used SpO_2_ values ranging from 85% to 100% and the corresponding range from a minimum S/F ratio 85 and ePFR 50 (representing a SpO_2_ of 85% on 100% FiO_2_) to a maximum S/F ratio 476 and ePFR 633 (representing a SpO_2_ of 100% on room air). To smoothen the ECDFs, we converted SpO_2_ from integer to continuous by adding uniformly distributed noise (+/- 0.5% with a maximum SpO_2_ of 100%). To calculate the rate of clinical deterioration at a particular level, we used a window centered at that level with width equal to one standard deviation (2.5 for SpO_2_, 100 for S/F ratio and 120 for ePFR). The dashed horizontal lines (Panels D-F) mark the rate of clinical deterioration in the entire dataset (1.85%).

In two important ways, this racial disparity was better revealed by ePFR and S/F ratio than by SpO_2_. First, the SpO_2_ distribution showed a narrower right-shift (K-S distance: 0.09 [UVA], 0.15 [Emory]; p < 0.01) than was revealed by the S/F ratio and ePFR distributions (K-S distance: 0.17 [UVA and Emory]; p < 0.01). Second, a racial influence on relationship between overt hypoxemia and outcomes (i.e. evidence of occult hypoxemia) was revealed much better by ePFR and S/F ratio than by SpO_2_. At UVA, when we modelled clinical deterioration using race, SpO_2_, and other baseline predictors, race was not found to be a significant predictor (p = 0.14). In contrast, when SpO_2_ was replaced by ePFR or S/F ratio in the model, race was a strong predictor (AOR 2.2-2.3, p < 0.01). Similarly, in the Emory data, race was a stronger predictor when clinical deterioration was modelled with ePFR or S/F ratio (AOR 1.20; p < 0.01) than with SpO_2_ (AOR 1.04; p < 0.01).

### 3.4) Sensitivity analyses at UVA

Repeating the analysis with restricted cubic splines for temperature, respiratory rate, heart rate, and mean arterial pressure did not significantly affect the predictive validity of the ePFR. When we extended the prediction horizon, the ePFR continued to outperform NEWS and SOFA (eFigure 3). The results were not meaningfully impacted by (a) excluding patients who died without transfer to ICU, (b) excluding patients who were discharged without outcome in < 24 hours (most likely to be ER visits and brief observation stays), and (c) varying of censoring time.

## Discussion

We studied how non-invasive measures of oxygenation inform on the clinical course of hospital patients with COVID-19. Our major findings are that a P/F ratio estimated by applying a model of the oxygen dissociation curve to pulse oximetry data (ePFR) had strong predictive validity for COVID-19 outcomes, and that pathological hypoxemia can be hidden in Black patients.

The adjusted odds ratio of 0.990-0.995 for a 1-point rise in ePFR reflects a strong relationship with clinical deterioration, considering the degree of variability that is typically observed in the ePFR (standard deviation around 120). It is equivalent to an odds ratio for deterioration of 1.7-2.7 for a 100-point decrease in the ePFR. On its own, ePFR outperformed complex multi-system dysfunction models like NEWS and Sepsis-3 in predicting deterioration. This likely reflects the uniqueness of COVID-19 as a syndrome in which acute deterioration occurs predominantly from impaired oxygenation. In syndromes like sepsis which consist of a multi-system organ dysfunction, the incorporation of ePFR into clinical criteria may enhance their performance.

As demonstrated in Figure 1, conventional markers of hypoxemia can be shown to have poor construct validity in common clinical scenarios. In some scenarios, these measures detect changes in hypoxemia when none exist. This may lead to false alarms and alarm fatigue. Even more concerning are the scenarios where these markers fail to sound early alarms about worsening hypoxemia. Such errors may lead to missed opportunities for early intervention and adverse patient outcomes. We found that the oxygenation measure ePFR, which combines information from SpO_2_ and oxygen flow, is less prone to these problems and may be the preferred alternative when measured P/F ratios are missing.

Importantly, this study validates the ePFR as a tool to demonstrate the real-world effects of racially biased pulse oximetry readings. We found no disparities in the probability of significant oxygen desaturation (such as SpO_2_ < 90), which suggests that clinicians were equitable in their efforts to prevent desaturation by adjusting supplemental oxygen. Yet, the separation in ePFR distributions was wide even at low values. This suggests that, on average, clinicians were achieving their SpO_2_ targets with lower supplemental oxygen settings in Black patients (eFigure 4). By itself, this finding could suggest that Black patients were hospitalized with less severe respiratory failure than others. But that conclusion is inconsistent with the finding that for comparable levels of oxygenation, Black patients were at higher risk of adverse outcomes than others (AOR 1.2-2.2). Together, these findings point to a phenomenon like occult hypoxemia which leads clinicians to use lower FiO_2_ settings because of a falsely reassuring SpO_2_ reading, leading to worse outcomes.

Our approach of comparing empirical cumulative distributions of ePFR is not limited by the need for arterial blood sampling. It will enable research into occult hypoxemia on a larger scale than has been possible to date. This new study design can equip consumers, advocates, politicians and regulators with evidence of racial disparities attributable to pulse oximetry to create the market forces and/or regulatory climate needed to bring an end to this important, longstanding source of structural inequity in healthcare. Until the time that pulse oximeter performance becomes racially equitable, the ePFR can be used to account for the influence of skin color on hypoxemia severity estimation.

The strength of our method for computing ePFRs is that it is grounded in the well-established physiology of the oxygen-hemoglobin dissociation curve. Unlike statistical imputation strategies (like multiple imputation), its reliability is not related to frequency of missing data. Additionally, our method lends itself to convenient implementation in large data sets including electronic medical records. Finally, the reproducibility of findings in diverse clinical settings is a major strength of this work.

A limitation of this work is the use of a care-delivery outcome. The reproducibility of results at diverse sites does improve confidence in findings. Still, several clinical practices may differ between sites and with times, affecting generalizability. Another limitation is our broad categorization of patients as Black or Non-Black. Skin color is not binary; skin color and racial identity are incongruous, and the race as recorded in the medical record is frequently misaligned with the patient’s racial identity^38^.

## Conclusions

P/F ratios estimated using the oxygen dissociation curve were simple to implement and accurately measured the severity of overt hypoxemic respiratory failure. In patients with COVID-19, they outperformed complex multi-system organ dysfunction models. Estimated P/F ratios may allow real-world modelling of racial disparities in outcomes attributable to occult hypoxemia from pulse oximetry.

## Take-home Points

### Study Question

Can we improve on the standard P/F ratio for oximetry-based detection of hypoxemia in COVID-19, especially in Black patients?

### Results

In this multicenter retrospective cohort study of 5319 hospital encounters for COVID-19, we found that a new, simple algorithm for non-invasive, oximetry-based estimation of the P/F ratio (P - partial pressure of arterial oxygen; F - fraction of inspired oxygen) outperformed other operational markers of hypoxemia in terms of availability, construct validity, predictive validity, and ability to characterize racial disparities.

### Interpretation

The P/F ratio estimated using the oxygen dissociation curve (ePFR) is an improved operational marker of hypoxemia for applications like clinical research, real time predictive modelling and post-marketing surveillance for bias in pulse oximetry devices.

## Data Availability

All data produced in the present study are available upon reasonable request to the authors

## Abbreviations

ABG: Arterial blood gas
AOR: Adjusted odds ratio
AUROC: Area under the receiver operating characteristics curve
CCI: Charlson comorbidity index
COVID-19: Novel coronavirus disease 2019
ECDF: Empirical cumulative distribution functions
ePFR: Estimated P/F Ratio
EUH: Emory University Hospital
EUH-M: Emory University Hospital Midtown
FiO_2_: Fraction of inspired oxygen
ICU: Intensive care unit
IRR: Inter-rater reliability
K-S: Kolmogorov-Smirnov
LPM: Liters per minute
NEWS: National Early Warning Score
PaO_2_: Partial pressure of arterial oxygen
P/F ratio: The ratio of the partial pressure of arterial oxygen to the fraction of inspired oxygen
S/F ratio: The ratio of the oxygen saturation from pulse oximetry to the fraction of inspired oxygen
SaO_2_: Arterial oxygen saturation
SARS-CoV-2: Severe acute respiratory syndrome coronavirus 2
SOFA: Sequential Organ Failure Assessment
SpO_2_: Oxygen saturation from pulse oximetry
TACS: Toward a COVID-19 Score
UVA: University of Virginia

## Acknowledgments

SMG (UVA) and RK (Emory) had full access to all of the data from their site used in the study and take responsibility for the integrity of the data and the accuracy of the data analysis.

All authors contributed substantially to the study design, data analysis and interpretation, and the writing of the manuscript.

This work was supported by the Manning fund for COVID-19 research. Dr. Moore is supported by the National Institutes of Health (U01AI150508). Dr. Holder is supported by the National Institutes of Health (K23GM37182), and has received speaker and consulting fees from Baxter International and Philips, respectively. Dr. Kamaleswaran was supported by National Institutes of Health (R01GM139967). Dr. Clark is an employee of Nihon Kohden Digital Health Solutions, Irvine CA. Dr. Moorman has equity in Medical Predictive Science Corporation, Charlottesville, VA, and consults for Nihon Kohden Digital Health Solutions, Irvine, CA, with proceeds donated to the University of Virginia Medical Foundation. For the remaining authors, no conflicts were declared.

## References

1. Gupta RK, Marks M, Samuels THA, et al. Systematic evaluation and external validation of 22 prognostic models among hospitalised adults with COVID-19: an observational cohort study. Eur Respir J 2020;56(6):2003498.

2. Wynants L, Calster BV, Collins GS, et al. Prediction models for diagnosis and prognosis of covid-19: systematic review and critical appraisal. BMJ 2020;369:m1328.

3. Knight SR, Ho A, Pius R, et al. Risk stratification of patients admitted to hospital with covid-19 using the ISARIC WHO Clinical Characterisation Protocol: development and validation of the 4C Mortality Score. BMJ 2020;370:m3339.

4. Liang W, Liang H, Ou L, et al. Development and Validation of a Clinical Risk Score to Predict the Occurrence of Critical Illness in Hospitalized Patients With COVID-19. JAMA Intern Med 2020;180(8):1081–1089.

5. Gerotziafas GT, Sergentanis TN, Voiriot G, et al. Derivation and Validation of a Predictive Score for Disease Worsening in Patients with COVID-19. Thromb Haemost 2020;120(12):1680–1690.

6. Saria S, Schulam P, Yeh BJ, et al. Development and Validation of ARC, a Model for Anticipating Acute Respiratory Failure in Coronavirus Disease 2019 Patients. Crit Care Explor 2021;3(6):e0441.

7. Fukuda Y, Tanaka A, Homma T, et al. Utility of SpO2/FiO2 ratio for acute hypoxemic respiratory failure with bilateral opacities in the ICU. PLOS ONE 2021;16(1):e0245927.

8. Singhal L, Garg Y, Yang P, et al. eARDS: A multi-center validation of an interpretable machine learning algorithm of early onset Acute Respiratory Distress Syndrome (ARDS) among critically ill adults with COVID-19. PLOS ONE 2021;16(9):e0257056.

9. Singh V, Kamaleswaran R, Chalfin D, et al. A deep learning approach for predicting severity of COVID-19 patients using a parsimonious set of laboratory markers. iScience 2021;24(12):103523.

10. Galloway JB, Norton S, Barker RD, et al. A clinical risk score to identify patients with COVID-19 at high risk of critical care admission or death: An observational cohort study. J Infect 2020;81(2):282–288.

11. Xie J, Hungerford D, Chen H, et al. Development and external validation of a prognostic multivariable model on admission for hospitalized patients with COVID-19 [Internet]. 2020 [cited 2021 Aug 13]. Available from: https://www.medrxiv.org/content/10.1101/2020.03.28.20045997v2

12. Vaid S, Kalantar R, Bhandari M. Deep learning COVID-19 detection bias: accuracy through artificial intelligence. Int Orthop 2020;44(8):1539–1542.

13. Guillamet MCV, Guillamet RV, Kramer AA, et al. Toward a Covid-19 Score-Risk Assessments and Registry [Internet]. 2020 [cited 2021 Aug 13]. Available from: https://www.medrxiv.org/content/10.1101/2020.04.15.20066860v1

14. Olsson T, Terent A, Lind L. Rapid Emergency Medicine score: a new prognostic tool for in-hospital mortality in nonsurgical emergency department patients. J Intern Med 2004;255(5):579–587.

15. Singer M, Deutschman CS, Seymour CW, et al. The Third International Consensus Definitions for Sepsis and Septic Shock (Sepsis-3). JAMA 2016;315(8):801–810.

16. Jakobsen JC, Gluud C, Wetterslev J, Winkel P. When and how should multiple imputation be used for handling missing data in randomised clinical trials – a practical guide with flowcharts. BMC Med Res Methodol 2017;17(1):162.

17. Seymour CW, Liu VX, Iwashyna TJ, et al. Assessment of Clinical Criteria for Sepsis: For the Third International Consensus Definitions for Sepsis and Septic Shock (Sepsis-3). JAMA 2016;315(8):762–774.

18. Gadrey SM, Clay R, Zimmet AN, et al. The Relationship Between Acuity of Organ Failure and Predictive Validity of Sepsis-3 Criteria. Crit Care Explor 2020;2(10):e0199.

19. Gadrey SM, Lau CE, Clay R, et al. Imputation of partial pressures of arterial oxygen using oximetry and its impact on sepsis diagnosis. Physiol Meas 2019;40(11):115008.

20. Hill AV. PROCEEDINGS OF THE PHYSIOLOGICAL SOCIETY: January 22, 1910. J Physiol 1910;40(suppl):iv–vii.

21. Goutelle S, Maurin M, Rougier F, et al. The Hill equation: a review of its capabilities in pharmacological modelling. Fundam Clin Pharmacol 2008;22(6):633–648.

22. Severinghaus JW. Simple, accurate equations for human blood O2 dissociation computations. J Appl Physiol 1979;46(3):599–602.

23. Jubran A, Tobin MJ. Reliability of pulse oximetry in titrating supplemental oxygen therapy in ventilator-dependent patients. Chest 1990;97(6):1420–1425.

24. Ebmeier SJ, Barker M, Bacon M, et al. A two centre observational study of simultaneous pulse oximetry and arterial oxygen saturation recordings in intensive care unit patients. Anaesth Intensive Care 2018;46(3):297–303.

25. Sjoding MW, Dickson RP, Iwashyna TJ, Gay SE, Valley TS. Racial Bias in Pulse Oximetry Measurement. N Engl J Med 2020;383(25):2477–2478.

26. Wong A-KI, Charpignon M, Kim H, et al. Analysis of Discrepancies Between Pulse Oximetry and Arterial Oxygen Saturation Measurements by Race and Ethnicity and Association With Organ Dysfunction and Mortality. JAMA Netw Open 2021;4(11):e2131674.

27. Vesoulis Z, Tims A, Lodhi H, Lalos N, Whitehead H. Racial discrepancy in pulse oximeter accuracy in preterm infants. J Perinatol 2021;1–7.

28. Henry NR, Hanson AC, Schulte PJ, et al. Disparities in Hypoxemia Detection by Pulse Oximetry Across Self-Identified Racial Groups and Associations With Clinical Outcomes*. Crit Care Med 2022;50(2):204–211.

29. Valbuena VSM, Barbaro RP, Claar D, et al. Racial Bias in Pulse Oximetry Measurement Among Patients About to Undergo Extracorporeal Membrane Oxygenation in 2019-2020: A Retrospective Cohort Study. CHEST 2022;161(4):971–978.

30. Burnett GW, Stannard B, Wax DB, et al. Self-reported Race/Ethnicity and Intraoperative Occult Hypoxemia: A Retrospective Cohort Study. Anesthesiology 2022;136(5):688–696.

31. Fawzy A, Wu TD, Wang K, et al. Racial and Ethnic Discrepancy in Pulse Oximetry and Delayed Identification of Treatment Eligibility Among Patients With COVID-19. JAMA Intern Med [Internet] 2022 [cited 2022 Jun 14];Available from: https://doi.org/10.1001/jamainternmed.2022.1906

32. Matt V, Matthew H. The retrospective chart review: important methodological considerations. J Educ Eval Health Prof [Internet] 2013 [cited 2019 Dec 14];10. Available from: https://www.ncbi.nlm.nih.gov/pmc/articles/PMC3853868/

33. Harris PA, Taylor R, Thielke R, Payne J, Gonzalez N, Conde JG. Research electronic data capture (REDCap)--a metadata-driven methodology and workflow process for providing translational research informatics support. J Biomed Inform 2009;42(2):377–381.

34. Harris PA, Taylor R, Minor BL, et al. The REDCap consortium: Building an international community of software platform partners. J Biomed Inform 2019;95:103208.

35. Krippendorff K. Estimating the reliability, systematic error and random error of interval data. Educ Psychol Meas 1970;30(1):61–70.

36. DeLong ER, DeLong DM, Clarke-Pearson DL. Comparing the areas under two or more correlated receiver operating characteristic curves: a nonparametric approach. Biometrics 1988;44(3):837–845.

37. R Core Team. R: A Language and Environment for Statistical Computing [Internet]. Vienna, Austria: R Foundation for Statistical Computing; 2018. Available from: https://www.R-project.org/

38. Klinger EV, Carlini SV, Gonzalez I, et al. Accuracy of Race, Ethnicity, and Language Preference in an Electronic Health Record. J Gen Intern Med 2015;30(6):719–723.

