## Supplemental Tables and Figures for "Overt and occult hypoxemia in patients hospitalized with novel coronavirus disease 2019"

**Appendix**

| eTable1: Separating pre-infection baseline SOFA from acute SOFA* at UVAMC | |
| --- | --- |
| **SOFA Component** | **Method of determining score** |
| Respiratory |  |
| Pre-infection baseline | We assumed a baseline partial pressure of arterial oxygen (PaO_2_) of at least 84 mmHg. This is the lowest integer value of assumed home PaO_2_ that assigns a baseline respiratory SOFA of 0 for patients that breathe room air (FiO_2_ of 0.21) at home.  Baseline supplemental oxygen settings were determined by chart review. From these, we estimated baseline fraction of inspired oxygen (FiO_2_) using standard clinical methods (Appendix Figure 1).  We divided the assumed home PaO_2_ (84mmHg) by the estimated home FiO_2_ to decide the baseline PF-ratio and respiratory SOFA scores.  In this way, we assigned a baseline respiratory SOFA of 0 to patients with no chronic oxygen use, 1 for oxygen use up to FiO_2_ of 0.28 (equivalent to 2 liters per minute by nasal cannula) and 2 for any oxygen use higher than that. |
| Maximal derangement | To determine acute maximal derangement in respiratory SOFA, we used both the imputed PaO_2_ values obtained by applying the Gadrey et al model of the oxygen dissociation curve to recorded oxygen saturation on pulse oximetry (SpO_2_).  We converted simultaneous recordings of supplemental oxygen settings to estimates of FiO_2_ using standard clinical methods (Appendix Figure 1).  To obtain the P-F ratio, we divided all PaO_2_ values by simultaneous FiO_2_ estimates. Based on these, we calculated the total respiratory SOFA at any given time.  We did not restrict the scores based on “respiratory support”. In other words, we allowed scores of 3 and 4 when ePFR was below 200 and 100 respectively, even though our patients were not mechanically ventilated. |
| Coagulation |  |
| Pre-infection baseline | We manually reviewed past laboratory results in the EMR and references in physician or nursing notes about known baseline derangements from verbal communications or faxed records. |
| Maximal derangement | We queried our data warehouse for platelet levels. In event of missing value, maximal derangement was assumed to be equal to pre-infection baseline value. |
| Hepatic |  |
| Pre-infection baseline | We manually reviewed past laboratory results in the EMR and references in physician or nursing notes about known baseline derangements from verbal communications or faxed records. |
| Maximal derangement | We queried our data warehouse for total bilirubin levels. In event of missing value, maximal derangement was assumed to be equal to pre-infection baseline value. |
| Renal |  |
| Pre-infection baseline | We manually reviewed past laboratory results in the EMR and references in physician or nursing notes about known baseline derangements from verbal communications or faxed records. |
| Maximal derangement | We queried our data warehouse for creatinine levels. In event of missing value, maximal derangement was assumed to be equal to pre-infection baseline value. Urine output was not used because of highly variable charting regularity. |
| Cardiovascular |  |
| Pre-infection baseline | We manually reviewed past laboratory results in the EMR and references in physician or nursing notes about known baseline derangements (like chronic hypotension requiring midodrine etc.) from verbal communications or faxed records.  The maximal baseline derangement was 1 as no patient with home infusions such as dobutamine or milrinone were a part of our cohort. |
| Maximal derangement | We queried our data warehouse for mean arterial pressures. The maximal derangement could only assume a value of 0 or 1 because vasopressor infusions are not initiated on acute care floors in our hospital. |
| Central Nervous System |  |
| Pre-infection baseline | We reviewed physician and nursing notes from current and past encounters to look for documentation of chronic mental state derangements. When Glasgow Coma Scale (GCS) was not charted, we looked for description of individual components of GCS. For example, “independent with activities of daily living and finances” was taken to mean a GCS of 15. “Occasionally gets confused at baseline” or mentions regarding inappropriate responses was taken to mean a GCS of 13-14. Incomprehensible speech or “non-verbal” baseline were taken to mean a GCS of 10-12. |
| Maximal derangement | We queried our data warehouse for nurse documented GCS levels. In event of missing value, maximal derangement was assumed to be equal to pre-infection baseline value. |
| * The acute rise in SOFA score was obtained by subtracting the maximally deranged value from pre-infection baseline value. This distinction was only available at UVA, not at EU | |

| eTable 2: Charlson Comorbidity Index | |
| --- | --- |
| Co-morbidity | Associated score |
| Myocardial Infacrtion  Congestive Heart Failure  Peripheral Vascular Disease  Cerebrovascular Disease  Dementia  Chronic Obstructive Pulmonary Disease  Connective Tissue Disease  Ulcer Disease  Mild Liver Disease  Diabetes | 1 |
| Hemiplegia  Moderate or severe renal disease  Diabetes with end organ damage  Any tumor  Leukemia  Lymphoma | 2 |
| Moderate or severe liver disease | 3 |
| Metastatic solid tumor  AIDS | 6 |

| eTable 3: Components of estimated P/F ratio, SOFA, and NEWS | | |
| --- | --- | --- |
| P/F ratio | SOFA | NEWS |
| Estimated PaO_2_ = $\left( \frac{23400}{\frac{1}{SpO_{2}}-0.99} \right)^{\frac{1}{3}}$ | Respiratory: 0 to 4^*^ based on estimated P/F ratio | Temperature: 0 to 3 |
|  | Cardiovascular: 0 to 1^†^ based on mean arterial pressure | Respiratory rate: 0 to 3 |
|  | Renal: 0 to 4 based on creatinine levels | Heart Rate: 0 to 3 |
| Estimated FiO_2_ calculations as described in Appendix Figure 1 | Coagulopathy: 0 to 4 based on platelet counts | Systolic Blood Pressure: 0 to 3 |
|  | Hepatic: 0 to 4 based on total bilirubin levels | Hypoxemia: 0 to 3 based on SpO_2_;  Add 2 points for any supplemental oxygen use |
|  | Neurologic: 0 to 4 based on recorded GCS level | Neurologic: 0 or 3 based on the AVPU score^‡^ |
| ^*^ We did not restrict the scores based on “respiratory support”. In other words, we allowed scores of 3 and 4 when ePFR was below 200 and 100 respectively, even though our patients were not mechanically ventilated.  ^†^ Patients can only get up 3 or 4 points if they are on vasopressors; since we censored prior to ICU transfer, none of our patients were on vasopressors.  ^‡^ The AVPU (Alert, Voice, Pain, Unresponsiveness) scale measures level of alertness. The NEWS model assigns 3 points for any rating other than “A” (alert). Since this scale is not routinely used in our institution, we assigned 3 points to any GCS rating lower than 15. | | |

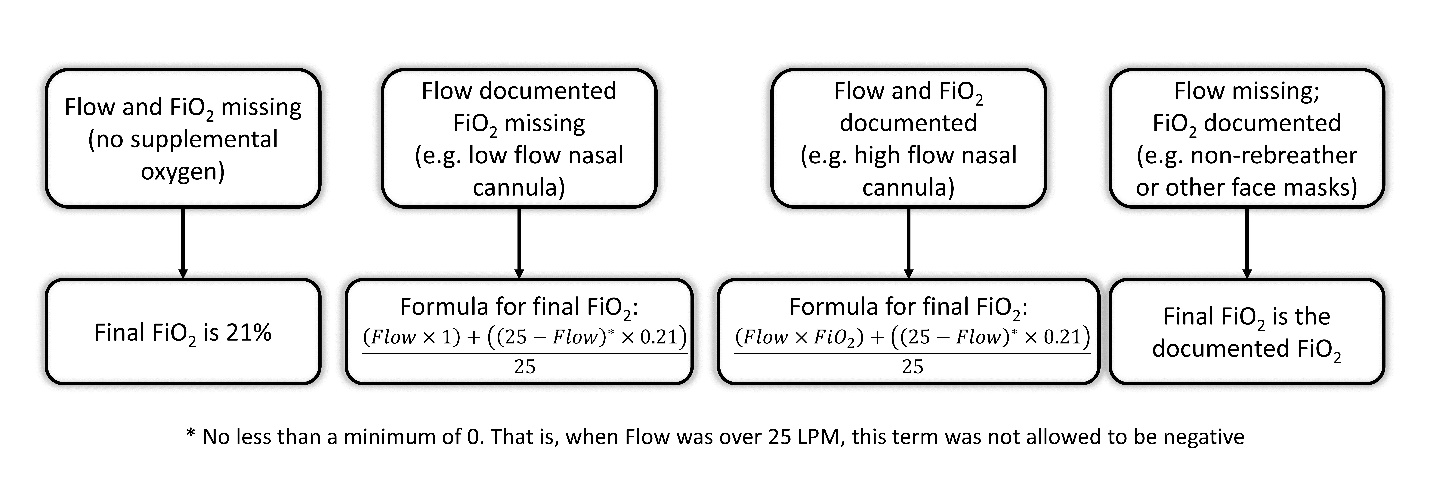

**eFigure 1:** This figure depicts our process of determining FiO_2_. It is designed to closely resemble how clinicians would determine FiO_2_ in their day-to-day clinical practice. It assumes an inspiratory flow rate of 25 liters per minute when accounting for entrainment of ambient air in low flow systems. (Abbreviations: Flow – supplemental oxygen flow rate; FiO_2_ – Fraction of Inspired Oxygen; LPM – Liters Per Minute)

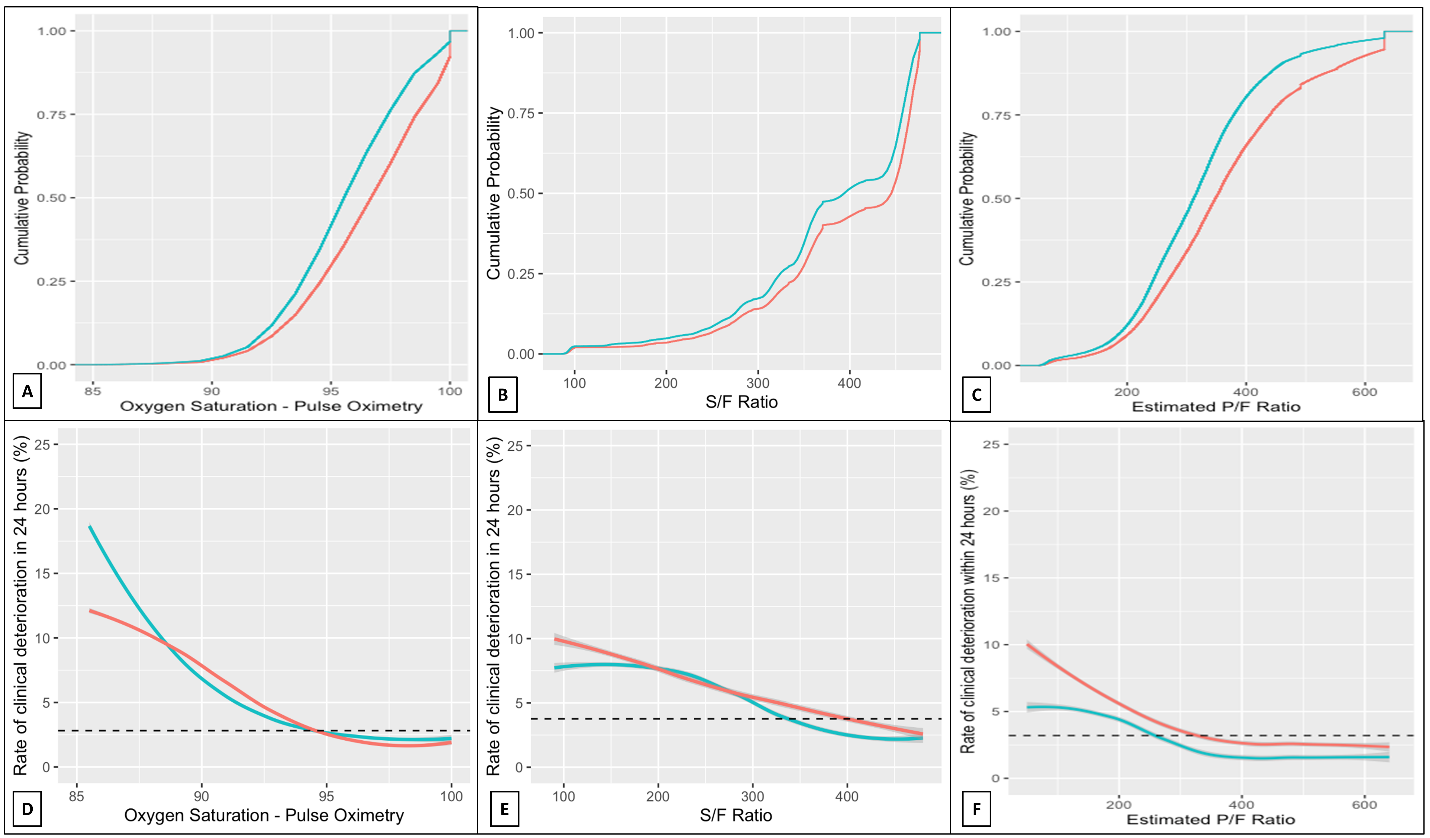
**eFigure 2:** Panels A - C show the Empirical Cumulative Distribution Functions for SpO_2_, S/F ratio, and ePFR respectively. This figure depicts the results from EU. Corresponding results from UVA are shown in Figure 3. Race is encoded by color (red - Black patients, blue - others). Similar to Figure 3, we observe a right shift in distributions of hypoxemia measures. By itself, this finding could suggest that Black patients were hospitalized with less intense respiratory failure than non-Black patients. But that conclusion is inconsistent with the finding that for comparable levels of oxygenation, Black patients were at higher risk of adverse outcomes than non-Blacks. Together, these findings point to a phenomenon like occult hypoxemia which leads clinicians to use lower FiO_2_ settings because of a falsely reassuring SpO_2_ reading, leading to worse outcomes. To make the plots directly comparable despite the varying scales of the hypoxemia measures, we used SpO_2_ values ranging from 85% to 100% and the corresponding range from a minimum S/F ratio 85 and ePFR 50 (representing a SpO_2_ of 85% on 100% FiO_2_) to a maximum S/F ratio 476 and ePFR 633 (representing a SpO_2_ of 100% on room air). To smoothen the ECDFs, we converted SpO_2_ from integer to continuous by adding uniformly distributed noise (+/- 0.5% with a maximum SpO_2_ of 100%). The dashed horizontal lines (Panels D-F) mark the rate of clinical deterioration in the entire dataset (2.9%).

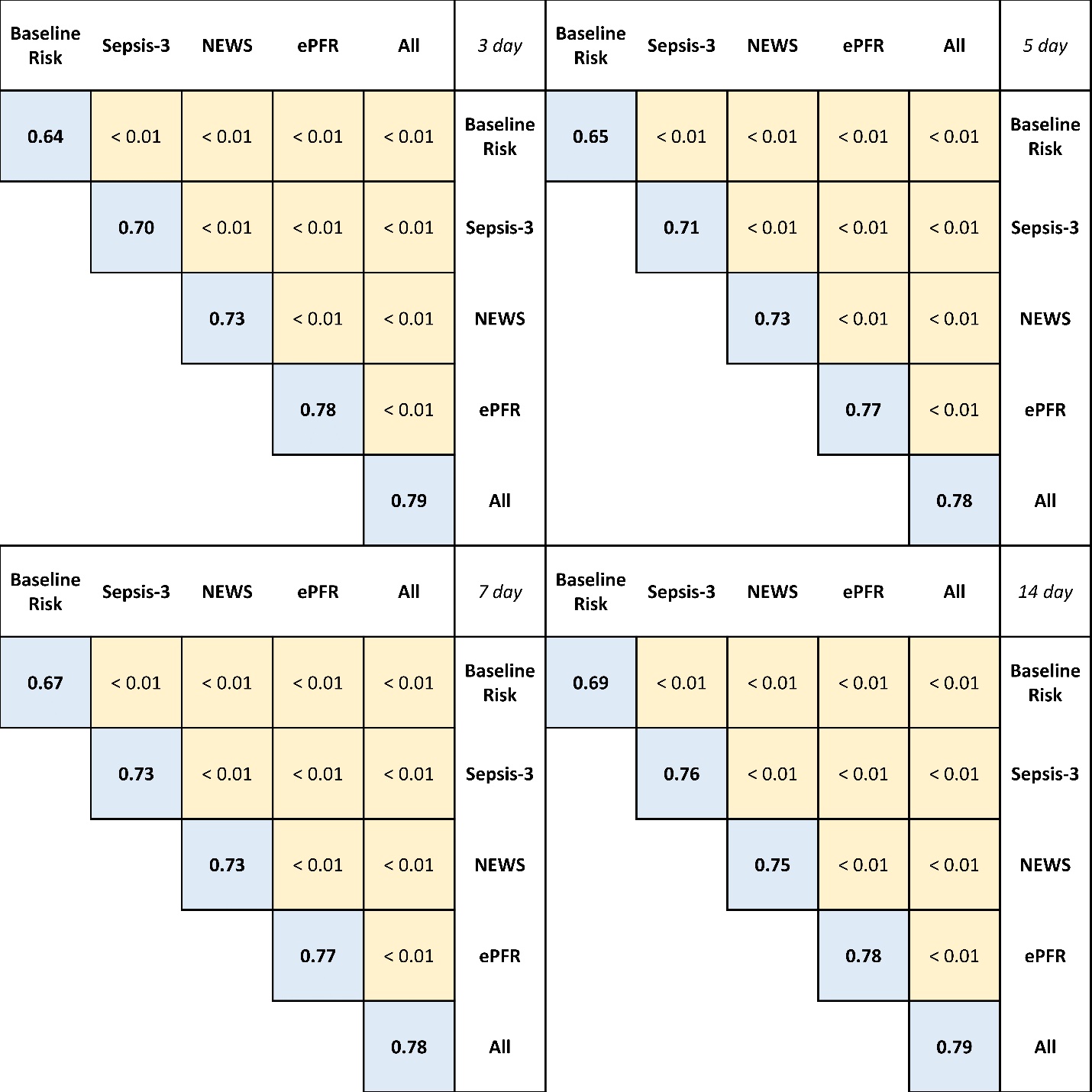

**eFigure 3:** For our primary analysis we used a prediction horizon of 24 hours (Figure 3). We repeated the analysis for 3 day, 5 day, 7 day, and 14 day horizons. The ePFR continued to outperform NEWS and Sepsis-3. However, the baseline risk model performed better with longer horizons (AUROC ranging from 0.63 for a 1 day horizon to 0.69 for a 14 day horizon).

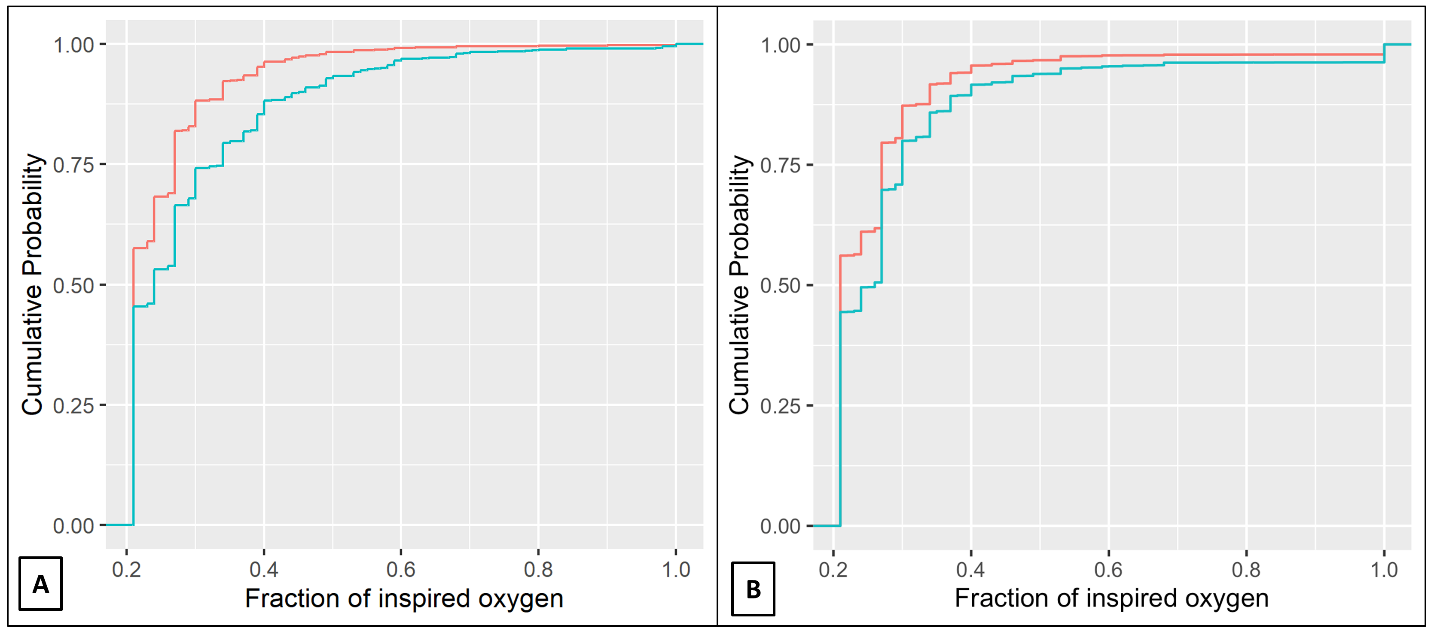
**eFigure 4:** The distribution of ePFR, that simultaneously account for oximetry values and clinicians’ response to it (supplemental oxygen adjustment), showed a wider racial disparity than was noted with the distribution of SpO_2_. This suggested that, on average, clinicians were achieving their SpO_2_ targets with lower supplemental oxygen settings in Black patients. This figure shows the empirical cumulative distribution functions for patients’ FiO_2_ at UVA (left) and Emory (right) which shows a left shift (lower FiO_2_) for Black patients (red) compared to non-Black patients (blue).
